# An ecological study of socioeconomic predictors in detection of COVID-19 cases across neighborhoods in New York City

**DOI:** 10.1101/2020.04.17.20069823

**Authors:** Richard S. Whittle, Ana Diaz-Artiles

**Affiliations:** Department of Aerospace Engineering, Texas A&M University, College Station, Texas, USA

## Abstract

**Background:** New York City was the first major urban center of the COVID-19 pandemic in the USA. Cases are clustered in the city, with certain neighborhoods experiencing more cases than others. We investigate whether potential socioeconomic factors can explain between-neighborhood variation in the number of detected COVID-19 cases.

**Methods:** Data were collected from 177 Zip Code Tabulation Areas (ZCTA) in New York City (99.9% of the population). We fit multiple Bayesian Besag-York-Mollié (BYM) mixed models using positive COVID-19 tests as the outcome and a set of 10 representative economic, demographic, and health-care associated ZCTA-level parameters as potential predictors. The BYM model includes both spatial and nonspatial random effects to account for clustering and overdispersion.

**Results:** Multiple different regression approaches indicated a consistent, statistically significant association between detected COVID-19 cases and dependent (under 18 or 65+ years old) population, male to female ratio, and median household income. In the final model, we found that an increase of only 1% in dependent population is associated with a 2.5% increase in detected COVID-19 cases (95% confidence interval (CI): 1.6% to 3.4%, *p* < 0.0005). An increase of 1 male per 100 females is associated with a 1.0% (95% CI: 0.6% to 1.5%, *p* < 0.0005) increases in detected cases. A decrease of $10,000 median household income is associated with a 2.5% (95% CI: 1.0% to 4.1% *p* = 0.002) increase in detected COVID-19 cases.

**Conclusions:** Our findings indicate associations between neighborhoods with a large dependent population, those with a high proportion of males, and low-income neighborhoods and detected COVID-19 cases. Given the elevated mortality in aging populations, the study highlights the importance of public health management during and after the current COVID-19 pandemic. Further work is warranted to fully understand the mechanisms by which these factors may have affected the number of detected cases, either in terms of the true number of cases or access to testing.

## Introduction

On January 21, 2020, the first case of Coronavirus disease 2019 (COVID-19) in the United States was reported in Washington State [1]. The first case was not reported in New York state until March 1, 2020 [2]. By the time the World Health Organization (WHO) declared a global pandemic on March 11, 2020, there were 345 cases in New York City (NYC), and this number skyrocketed to nearly 18,000 cases just two weeks later [2, 3]. NYC rapidly became the epicenter of the pandemic in the United States, with a transmission rate five times higher than the rest of the country, and over a third of all confirmed national cases by early April [4].

During a pandemic, there is likely to be large variation in both disease transmission and disease testing between regions [5]. These two factors cause large variation in disease reporting between different areas [6]. This is particularly true in the early stages of the outbreak, before disease testing has become widespread and standardized.

Contemporary and historical studies on previous pandemics, including H1N1 pandemics in 1918 and 2009, suggest that socioeconomic factors on a national level can affect detection rates and medical outcomes [7–9]. Thus, socioeconomic factors such as young or old populations, race, affluence, inequality, poverty, unemployment, insurance, or access to healthcare may account for differences in reported cases of COVID-19 between neighborhoods in NYC. The aim of this ecological study was to identify potential neighborhood-level socioeconomic determinants of the number of detected COVID-19 cases and explain between-neighborhood variation during the early, exponential growth stage of the pandemic in NYC: from the first detected case in March 1 until April 5, 2020.

## Materials and methods

### Data collection

Data on positive COVID-19 cases were collected from NYC Department of Health and Mental Hygiene (DOHMH) Incident Command System for COVID-19 Response (Surveillance and Epidemiology Branch in collaboration with Public Information Office Branch) [2]. Since the NYC DOHMH was discouraging people with mild to moderate symptoms from being tested during the time period covered, the data primarily represents people with more severe illness. Since at the time of writing the pandemic is still ongoing, data were taken at a snapshot on April 5, 2020. This date was chosen to cover the first month of the pandemic in NYC, since understanding early etiology of the pandemic and local influences is important in helping to inform future management [10]. Data were a cumulative count up to and including April 5, 2020. On this date, NYC had a cumulative total of 64,955 cases [11], including deaths and hospitalizations.

The available dataset included 64,512 cases (99.3% of total cases), with each case representing a positive diagnosis of COVID-19 along with the patient’s Zip Code Tabulation Area (ZCTA). ZCTAs are generalized areal representations of United States Postal Service (USPS) Zip Code service areas. ZCTAs were the areas in which patients reported their home address, as opposed to either where they became symptomatic or where they reported for testing/treatment. The area of interest covered 177 ZCTAs within NYC, from 10001 (Chelsea, Manhattan) to 11697 (Breezy Point, Queens). Of these cases, there were 4,712 where the patient ZCTA was unknown and thus, these cases were discarded, leaving 59,800 cases (92.1% of total cases). Note that this total is not meant to be an indicator of the total number of COVID-19 cases at this time, rather the count of *detected* cases. Fig 1A shows a histogram of cases by ZCTA as at April 5, 2020, grouped by the five boroughs of NYC (Bronx, Brooklyn, Manhattan, Queens, and Staten Island); Fig 1B displays these cases on a map as a percentage of total ZCTA population.

**Fig 1.**
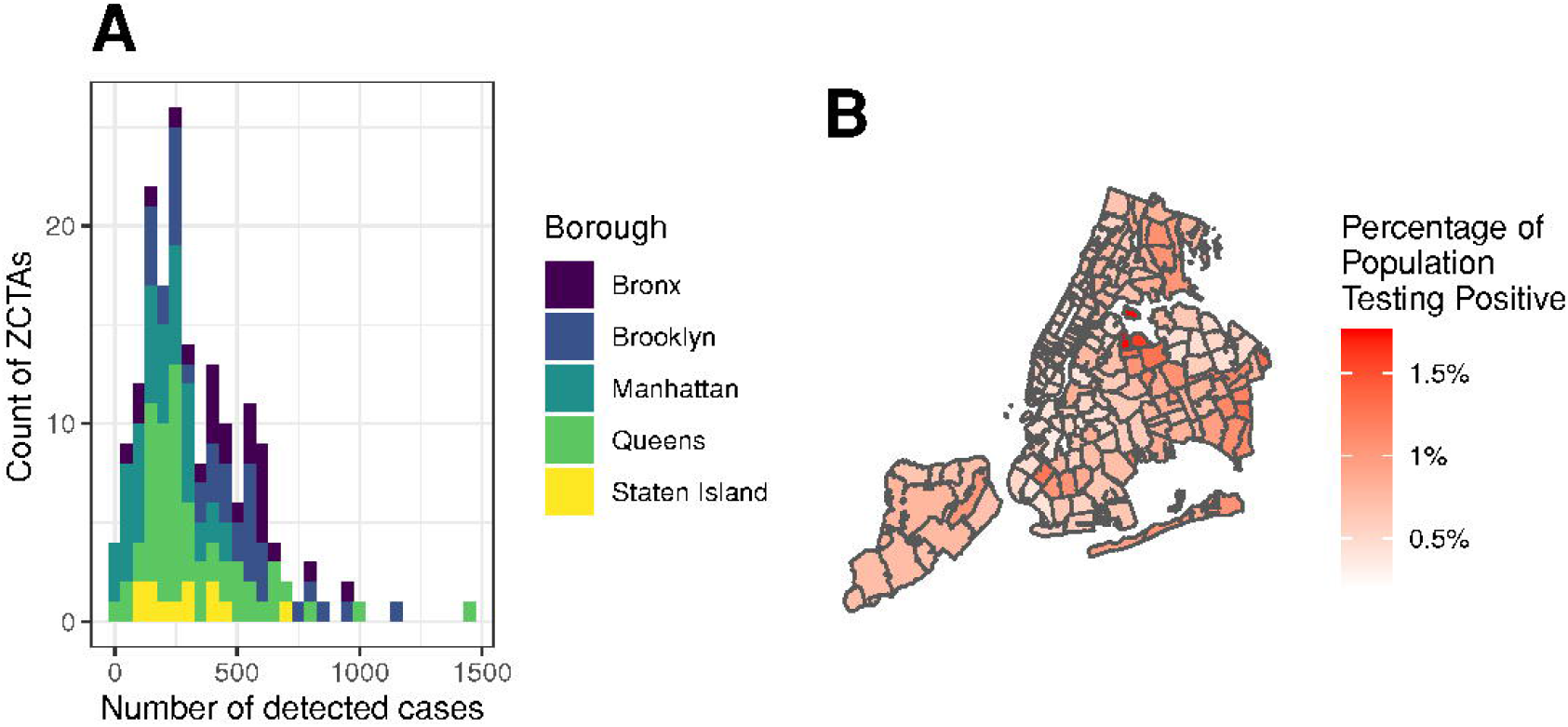
New York City detected COVID-19 cases by Zip Code Tabulation Area (ZCTA). As at April 5, 2020. A: Histogram of cases by ZCTA, grouped by borough. B: Detected cases as a percentage of total population.

Data on potential predictor variables were collected from the United States Census Bureau American Community Survey (ACS). ACS is a continuous sample survey of 3.5 million households every year including questions beyond the decadal census on subjects such as education, employment, internet access, and transportation. Data were collected at ZCTA level from the ACS 2014-2018 5-year estimate [12], which is the most recent publicly available. The 5-year estimate was chosen instead of the most recent 1-year estimate because the latter was not available in an aggregated form at ZCTA level. Whilst the 5-year estimate is less current, it has a smaller margin of error than the 1-year estimates and greater statistical reliability for small geographic areas.

### Demographic parameters

Four demographic parameters were included in the study: Percentage of dependent population, *Dependents*; percentage of aged population, *Aged*; males per 100 females, *MFR*; and percentage of the population identifying as white, Race. Dependent population was defined as the percentage of the total population aged under 18 or 65+ i.e., populations typically economically inactive. Aged population was the percentage of the total population 65+. The increased severity of COVID-19 with increasing age has been well documented [13], and there has been recent evidence of asymptomatic carrier transmission particularly amongst young people [14, 15]. Males per 100 females was chosen to capture the balance of sex in the population. We were interested in whether sex differences lead to significant variation in detected cases. Finally, some reports suggest a racial disparity in case detection rates across the USA. A report from NYU Furman Center for housing, neighborhoods, and urban policy suggests mortality rates are higher amongst the city’s “Hispanic, Black, and non-Hispanic/Latino: Other” populations [16]. For the present study, we chose to include the percentage of the population that identify as white (alone or in combination with another race) as a combined indicator of all minority populations. The distributions of demographic predictors in the area of interest are shown in Fig 2.

**Fig 2.**
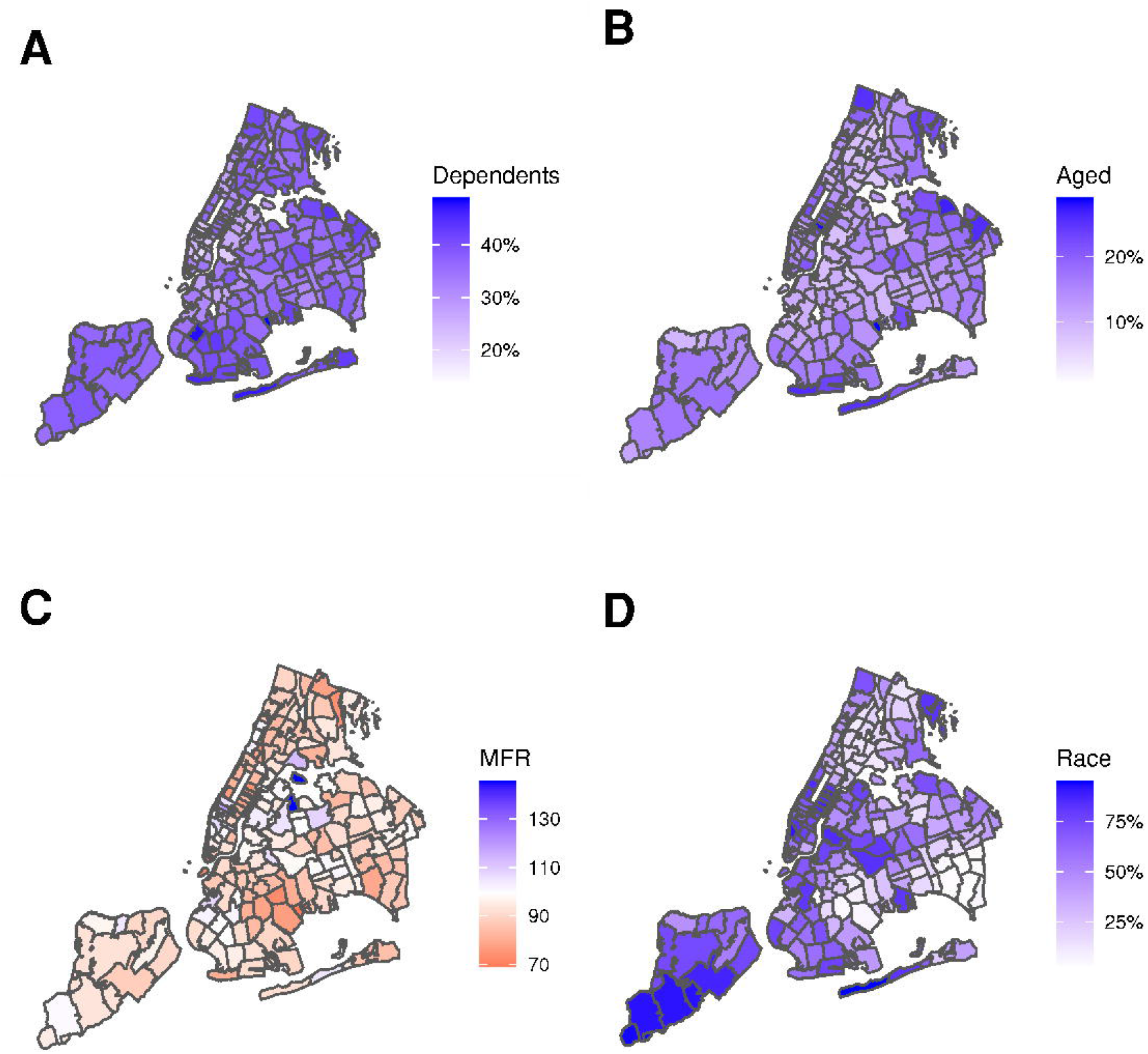
New York City demographic predictors by Zip Code Tabulation Area (ZCTA). Data based on American Community Survey (ACS) 2018 5-year estimates. A: *Dependents*, percentage of population aged under 18 or 65+. B: *Aged*, percentage of population aged 65+. C: *MFR*, males per 100 females. D: *Race*, percentage of population that identify as white (alone or in combination with another race).

### Economic parameters

Four economic parameters were included in the study: Gini index, *Gini*; median household income, *Income*; Percentage of labor force unemployed, *Unemployment*; and percentage of population living below the poverty threshold, *Poverty*. Gini index is a measure of economic inequality ranging from 0 to 1. An index of 0 indicates all the wealth in an area is divided equally amongst the population, whilst an index of 1 indicates all the wealth is held by one individual. Whilst some studies have argued against the adverse effects of unequal income [17], an association has been demonstrated between inequality and population health [18]. We also included household income, which was a significant predictor for hospitalizations in the 2009 influenza pandemic [19]. Specifically, in the present study we use median household income as a ZCTA-level predictor. Finally, unemployment and poverty both have documented association with health outcomes, including in pandemic scenarios [20, 21]. Whilst there is some level of collinearity between these two variables, we include both as one relates to the economically active labor force whereas the other to the total population. The distribution of economic predictors in the area of interest are shown in Fig 3.

**Fig 3.**
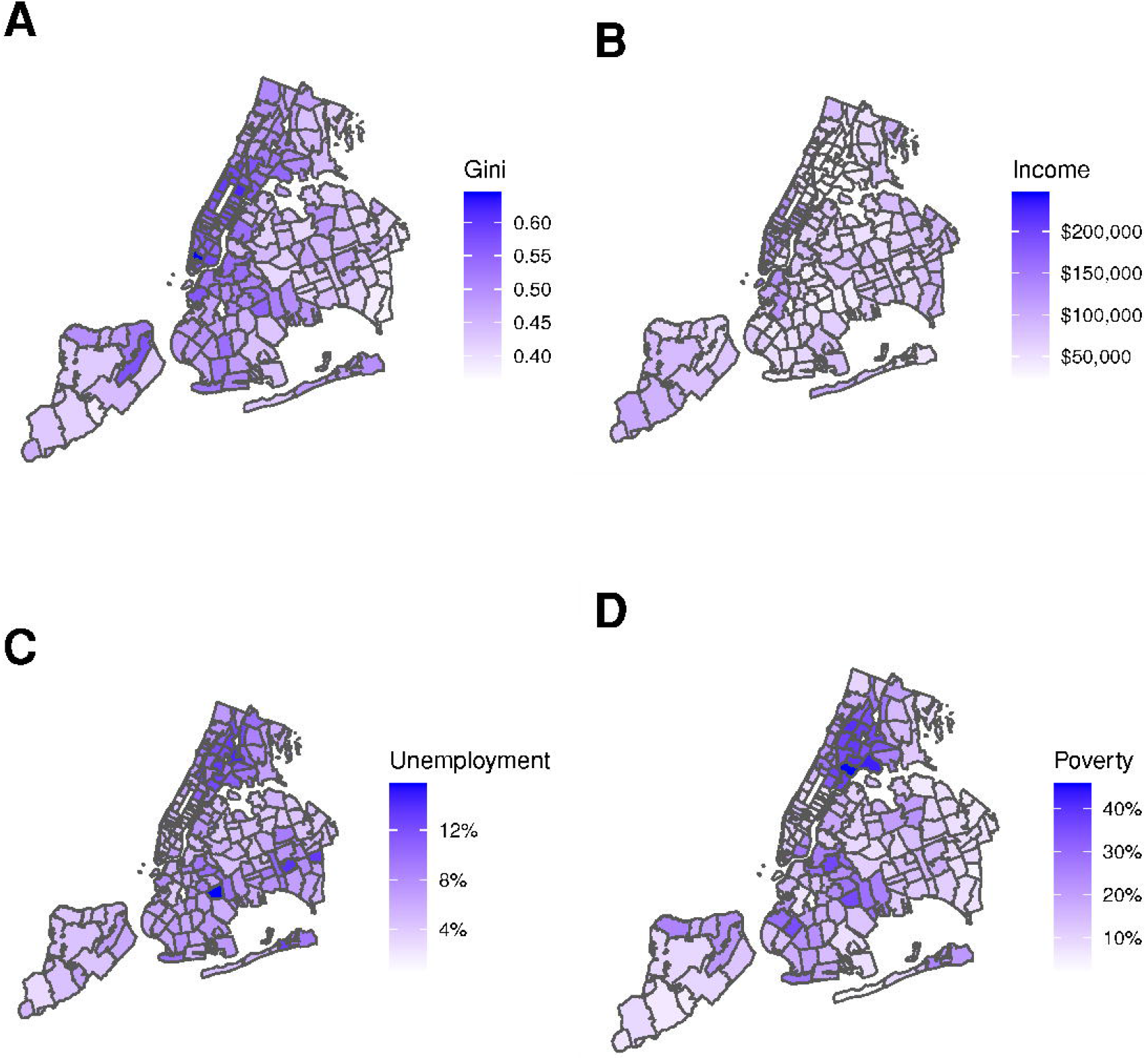
New York City economic predictors by Zip Code Tabulation Area (ZCTA). Data based on American Community Survey (ACS) 2018 5-year estimates. A: *Gini*, Gini index. B: *Income*, median household income. C: *Unemployment*, percentage of working age population unemployed. D: *Poverty*, percentage of total population living below the poverty threshold.

### Health parameters

Two parameters related to healthcare access were included in the study: percentage of population uninsured, *Uninsured*; and total number of hospital bed per 1000 people within 5 km, *Beds*. It has been documented that lack of insurance can delay access to timely healthcare, particularly during pandemics [22]. We hypothesized that this could affect virus transmission and/or access to testing, therefore affecting detection rates. Finally, we chose *Beds* as a parameter related to proximity to healthcare, which has been shown to be inversely associated with adverse outcomes in other geospatial public health studies [23]. For a city containing multiple hospitals such as NYC, we defined a proximity metric in this study as population normalized number of hospital beds within 5 km. The distribution of health related predictors in the area of interest are shown in Fig 4A and Fig 4B. Fig 4 also shows two other factors used in the model; Fig 4C shows the population of each ZCTA used as the model exposure, and Fig 4D shows the neighborhood connectivity between ZCTAs, used for spatial effects.

**Fig 4.**
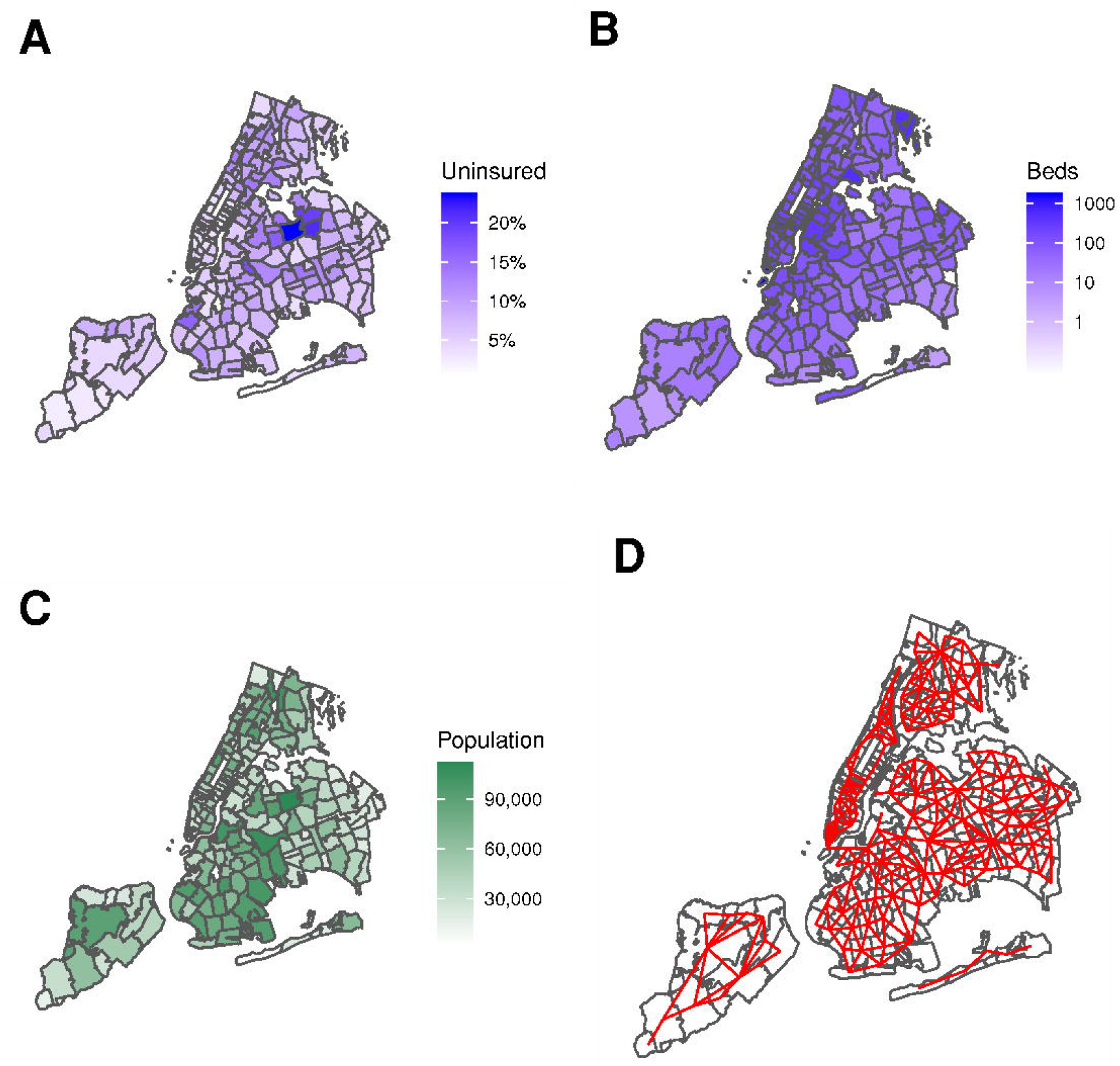
New York City health predictors by Zip Code Tabulation Area (ZCTA). Data based on American Community Survey (ACS) 2018 5-year estimates. A: *Uninsured*, percentage of total population uninsured. B: *Beds*, total number of hospital beds per 1000 people within 5 km. C: Population (exposure). D: Neighborhood connectivity.

### Statistical analysis

#### Base model

Prior to analysis of potential predictors, we considered multiple base regression models. Given the significant spatial correlation in the present case data as evidenced by the Moran Index, *I* (176) = 0.481, *p* < 0.0005 [24], we explored potential regression models both with and without spatial effects. We compared four base models (no predictors): 1) a Poisson model with random intercept; 2) a Poisson Besag-York-Mollié (BYM) model [25]; 3) a Negative Binomial model with random intercept; and 4) a Negative Binomial BYM model. The BYM model is the union of a Besag model [26], *υ*, and a nonspatial random effect, *ν*, such that the linear predictor for spatial unit *i, η*_*i*_, is given by Eq 1:

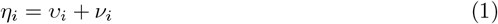

where *υ*_*i*_ has an intrinsic conditional autoregressive (ICAR) structure [27]. We used the reparameterization of the BYM model proposed by Riebler et al. [28], known as the BYM2 model and shown in Eq 2:

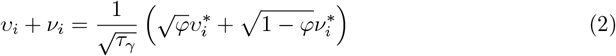

where *τ*_*γ*_ is the overall precision hyperparameter, *φ*∈ [0, 1] is the mixing hyperparameter representing the proportional division of variance between the spatial and nonspatial effects, *υ*^*\**^ is the spatial (ICAR) effect with a scaling factor such that Var (*υ*^*\**^) *≈* 1, and *ν*^*\**^ is the nonspatial random-effect with *ν*^*\**^ *∼* N (0, 1). Penalized complexity (PC) priors are applied to hyperparameters *τ*_*γ*_ and *φ* (compared to log-gamma priors in the random intercept model) [29]. All four models used ZCTA population as the exposure and a log-link function. We selected the model with the lowest Deviance Information Criterion (DIC) [30], representing the best trade-off between model fit and complexity.

Characteristics for the four base models examined, including hyperparameters, are shown in Table 1. The two Poisson models both had significantly lower DIC than the Negative Binomial models. The Poisson BYM2 model (Model 2) was marginally better than the simple random effect model. The Poisson BYM2 model was used for all future analyses and regressions.

**Table 1.** Characteristics of four different base models (no predictors). Lower Deviance Information Criterion (DIC) represents a better trade off between model fit and complexity. Models 1 and 3 have a random intercept, models 2 and 4 follow a BYM2 structure. 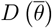, deviance of mean model parameters *θ*; *p*_*D*_, effective number of parameters.

| Model | Distribution | Parameters | Hyperparameters | $D(\bar{\theta})$ | $p_D$ | DIC |
| --- | --- | --- | --- | --- | --- | --- |
| Model 1* | Poisson | $\beta_0, \nu_i$ | $\tau_\nu$ | 1319.89 | 168.76 | 1657.41 |
| Model 2** | Poisson | $\beta_0, v_i^*, \nu_i^*$ | $\tau_\gamma, \varphi$ | 1320.75 | 166.88 | 1654.51 |
| Model 3† | Negative Binomial | $\beta_0, \nu_i$ | $n, \tau_\nu$ | 2096.86 | 3.00 | 2102.86 |
| Model 4‡ | Negative Binomial | $\beta_0, v_i^*, \nu_i^*$ | $n, \tau_\gamma, \varphi$ | 1646.63 | 124.74 | 1896.11 |

Notes:
\*Model 1: $y_i | \lambda_i \sim \text{Pois}(\lambda_i)$ , $\log(\lambda_i) = \eta_i + \log(E_i) = \beta_0 + \nu_i + \log(E_i)$
\*\*Model 2: $y_i | \lambda_i \sim \text{Pois}(\lambda_i)$ , $\log(\lambda_i) = \eta_i + \log(E_i) = \beta_0 + \frac{1}{\sqrt{\tau_\gamma}} (\sqrt{\varphi} v_i^* + \sqrt{1 - \varphi} \nu_i^*) + \log(E_i)$
†Model 3: $y_i | \lambda_i \sim \text{NegBin}(n, \lambda_i)$ , $\log(\lambda_i) = \eta_i + \log(E_i) = \beta_0 + \nu_i + \log(E_i)$
‡Model 4: $y_i | \lambda_i \sim \text{NegBin}(n, \lambda_i)$ , $\log(\lambda_i) = \eta_i + \log(E_i) = \beta_0 + \frac{1}{\sqrt{\tau_\gamma}} (\sqrt{\varphi} v_i^* + \sqrt{1 - \varphi} \nu_i^*) + \log(E_i)$
Symbols: $y_i$ , count of cases in Zip Code Tabulation Area (ZCTA) $i$ ; $\lambda_i$ , expected cases in ZCTA $i$ ; $E_i$ , population in ZCTA $i$ ; $\eta_i$ , linear predictor for ZCTA $i$ ; $\beta_0$ , intercept; $\nu_i$ , nonspatial random-effect; $\nu_i^*$ , scaled nonspatial random-effect; $v_i^*$ , scaled spatial random-effect with intrinsic conditional autoregressive structure; $\tau_\nu$ , precision for nonspatial random effect, log-Gamma prior; $\tau_\gamma$ , overall precision, penalized complexity (PC) prior; $\varphi$ , mixing parameter, PC prior; $n$ , overdispersion parameter, PC Gamma prior.

#### Adding predictors

Multiple regression models were built using a method adjusted from Nikolopoulos et al. [31]. In the univariable models, we considered each predictor variable separately (i.e., one model per variable). In the multivariable model we considered all predictor variables together. We further built a partial multivariable model using only those predictors that were significant in the univariable models. Finally we built a model using stepwise backwards elimination procedure, starting with the fully saturated model and removing the least significant predictor until we were left with a model containing only significant predictors [31]. In all cases, the expected number of detected COVID-19 cases in ZCTA *i, λ*_*i*_, was represented by Eq 3:

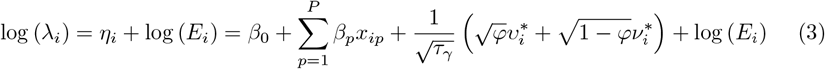

where *E*_*i*_ is the exposure (i.e., population) for ZCTA *i, β*_0_ is the intercept, *β*_*p*_ is coefficient of the fixed effect for predictor *p* ∈{1…*P*}, *x*_*ip*_ is the value of predictor *p* in ZCTA *i*, and the spatial and nonspatial random effects for ZCTA *i* are described by the BYM2 model detailed above. Vague Gaussian priors are assumed on all *β*.

#### Model fitting

Regression estimates are presented as mean and 95% confidence intervals (CI) sampled from the posterior marginal distribution, along with corresponding p-values. We used posterior tail-area of the fixed effects as a Bayesian counterpart to p-value [32]. All significance levels were two-sided with p-value of < 0.05 considered statistically significant. Statistical analysis was performed using R Statistical Software (version 3.6.2; R Foundation for Statistical Computing, Vienna, Austria). Models were fit via integrated nested Laplace approximation [33] using the R-INLA package [34]. Vague priors were assumed on all models.

## Results

As at April 5, 2020, 59,800 COVID-19 cases were reported with a known ZCTA. The highest number of cases in any particular ZCTA was 1,446 in ZCTA 11368 (Corona, Queens), whilst the lowest was 7 in ZCTA 10006 (Wall St, Manhattan). With respect to the proportion of the population, ZCTA 10006 also had the lowest detection rate (0.20%). The highest detection rate was 1.76% in ZCTA 11370 (East Elmhurst, Queens). This ZCTA also includes Rikers Island. On average, 0.71% of the total NYC population had tested positive for COVID-19.

### Base model

Using the base model, Fig 5A shows the area specific relative risk *ζ*_*i*_. A value of *ζ*_*i*_ = 1 represents detected cases in line with the total population average (0.71% of the population of area *i* has detected positive), whilst, for example, a value of *ζ*_*i*_ = 2 represents detected cases twice the total population average (1.42%). Fig 5B shows the posterior probability that the relative risk is greater than 1, *p* (*ζ*_*i*_ *>* 1 | **y**). The map shows that the highest risk area is East Elmhurst, Queens, including Rikers Island, with three other significant clusters in East Bronx, Southeast Queens, and Southwest Brooklyn.

**Fig 5.**
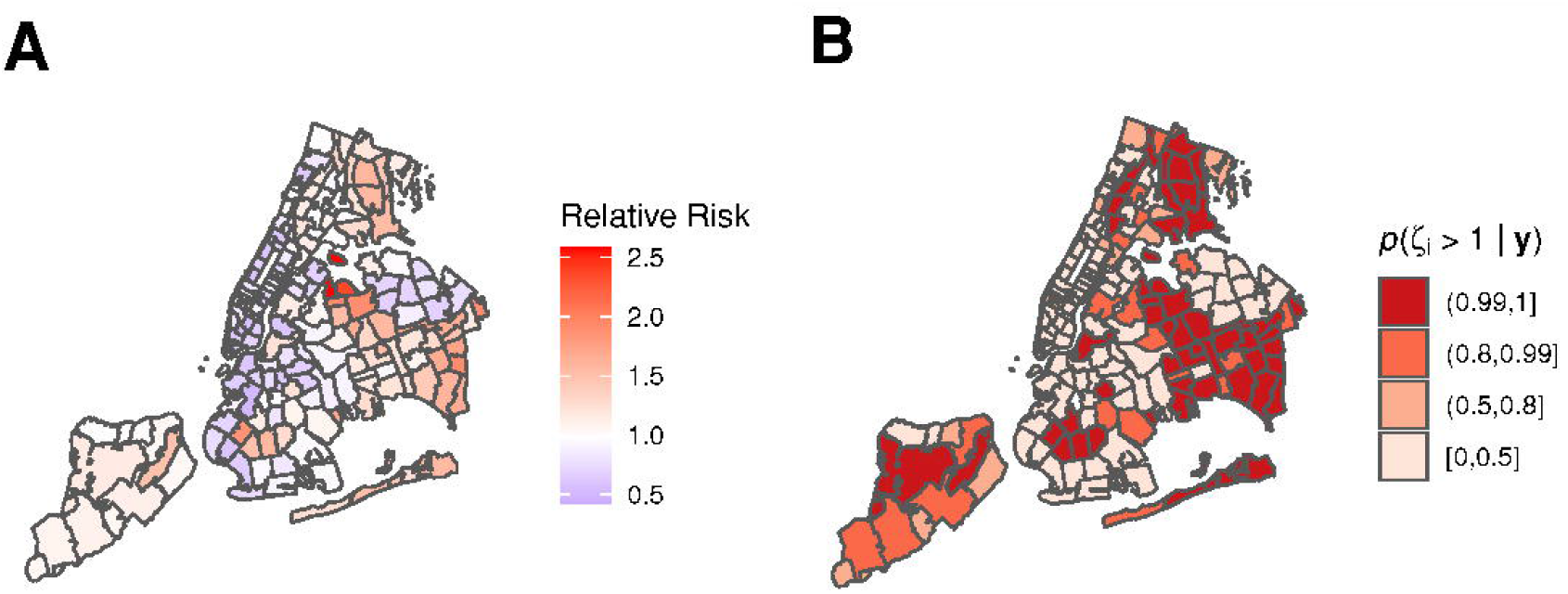
Disease mapping model for COVID-19 cases in New York City by Zip Code Tabulation Area (ZCTA). As at April 5, 2020, using base Poisson BYM2 model with no predictors. The area specific relative risk is multiplied by the total population average COVID-19 detection rate (0.71%) to give the area specific detection rate. A: Area specific relative risk, *ζ*_*i*_. B: Posterior probability for relative risk, *p* (*ζ*_*i*_ *>* 1|**y**).

### Adding predictors

Spread and collinearity of the predictors was assessed through histograms, bivariate scatterplots, and Pearson correlation coefficients. The strongest collinearities existed between income, poverty, and unemployment. There was only one bivariate correlation above 0.7 (median household income and poverty), and none above 0.8. It was decided to leave all predictors in the analysis and to build multiple regression models in order to consider the effects of collinearity. Fig 6 shows panel plots of the bivariate relations between the predictors.

**Fig 6.**
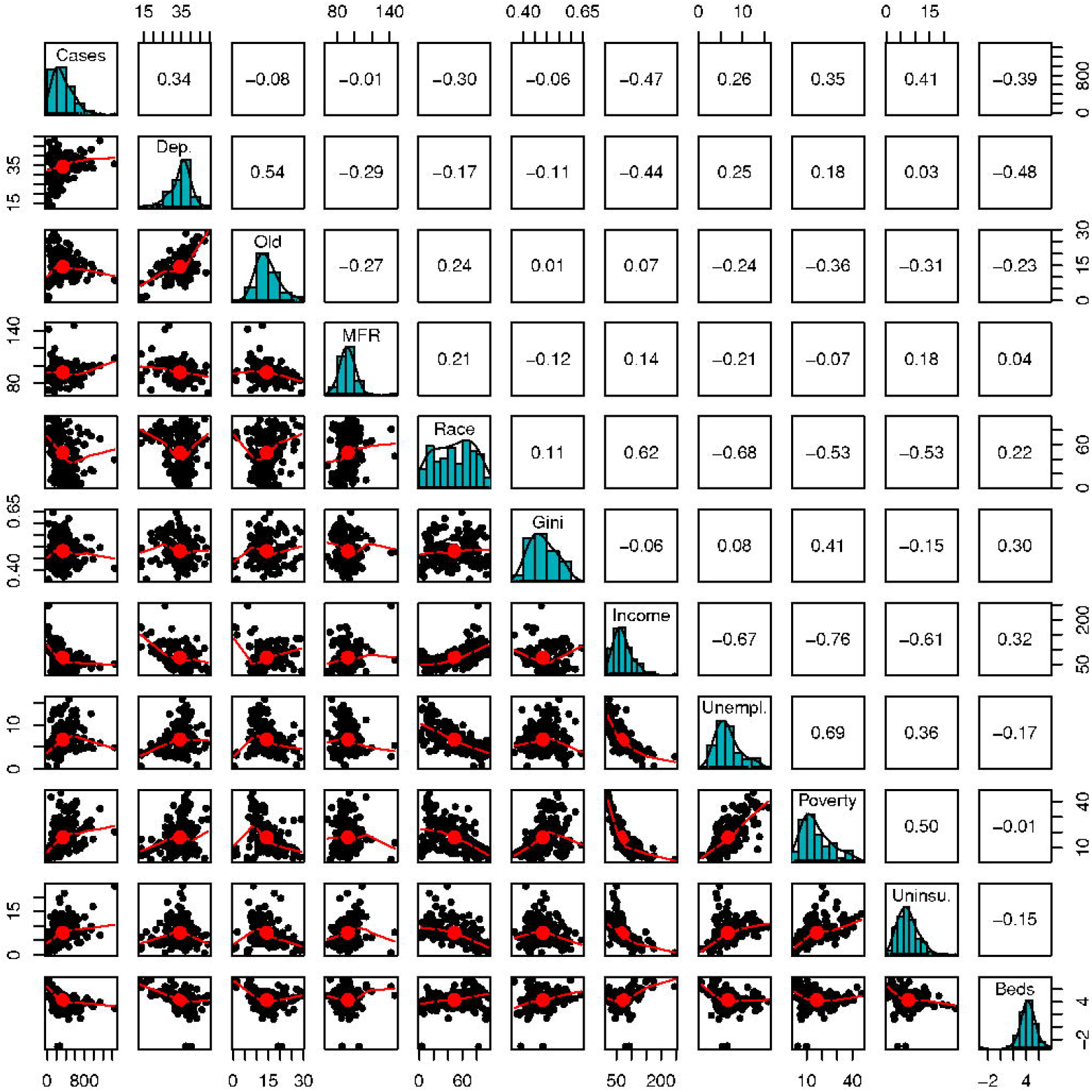
Panel plot showing bivariate relationships between predictors. Diagonal: Distributions of cases and all 10 predictor variables. Lower: Bivariate scatter plots. Upper: Pearson correlations between pairs of predictors.

Table 2 shows a summary of the regression estimates from the different regression models investigated. In particular, two predictors appear in all four models: percentage of dependent population and median household income. Dependency is significant in all four models, whilst income is significant in three of them. Percentage change in the detected COVID-19 cases per unit change in the predictors can be found from exp (*β*).

**Table 2.**
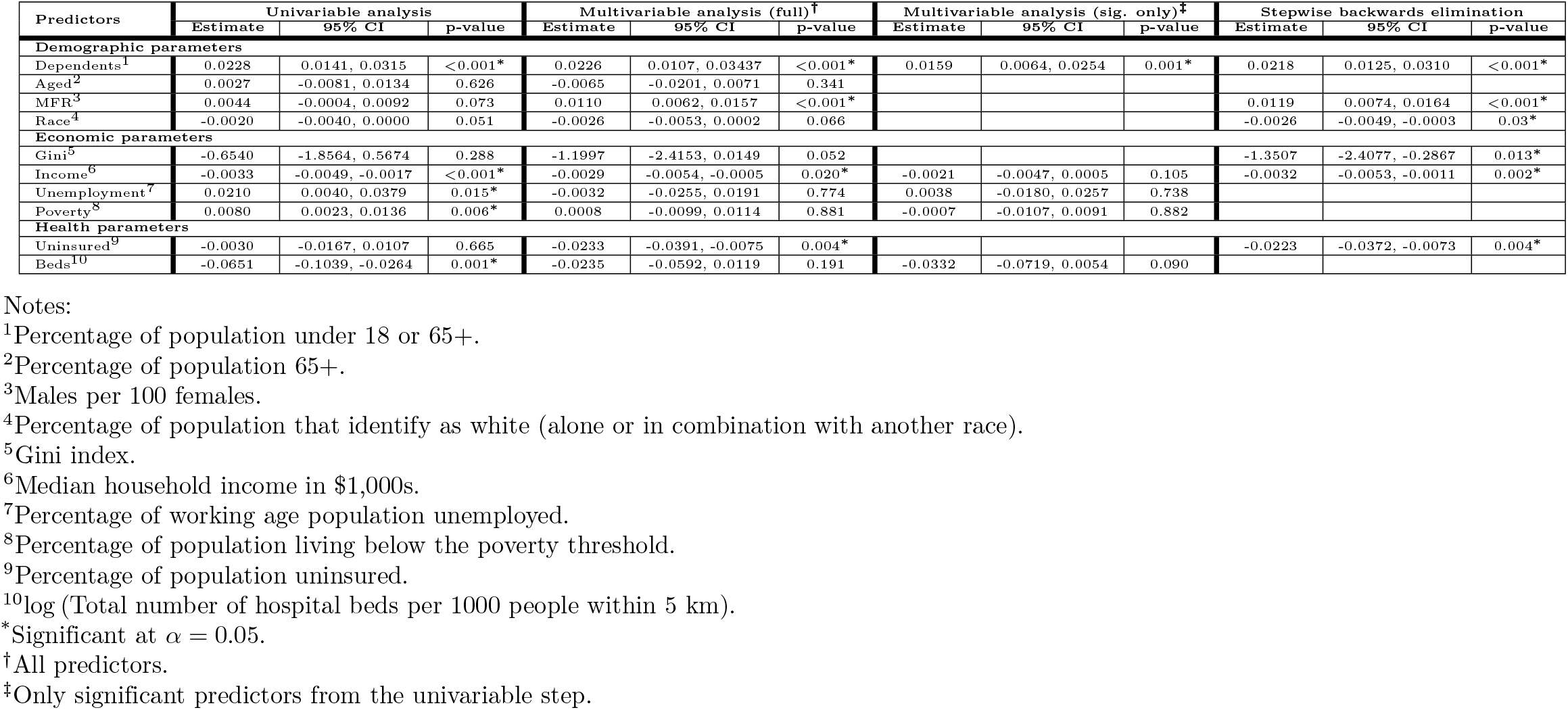
Regression estimates for association of Zip Code Tabulation Area (ZCTA) level predictors with detected COVID-19 cases in New York City as at April 5, 2020.

Concerning dependency, a 1% increase in the percentage of dependent population leads to an increase in detected COVID-19 cases of 2.3% (95% CI: 1.4% to 3.2%, *p* < 0.0005) in the univariable model, an increase of 2.3% (95% CI: 1.1% to 3.5%, *p* < 0.0005) in the full multivariable model, an increase of 1.6% (95% CI: 0.6% to 2.6%, *p* = 0.001) in the partial multivariable model, and an increase of 2.2% (95% CI: 1.3% to %, *p* < 0.0005) in the stepwise backwards elimination model. Concerning income, a $10,000 decrease in median household income leads to an increase in COVID-19 cases of % (95% CI: 1.7% to 5.0%, *p* < 0.0005) in the univariable model, an increase of 2.9% (95% CI: 0.5% to 5.5%, *p* = 0.020) in the full multivariable model, and an increase of 3.2% (95% CI: 1.1% to 5.4%, *p* = 0.003) in the stepwise backwards elimination model. Income was insignificant in the partial multivariable model (2.2%, 95% CI: -0.5% to 4.8%, *p* = 0.106).

Males per 100 females (MFR) and race do not appear in the partial multivariable model since their univariable predictors were marginally insignificant (*p* = 0.073 and *p* = 0.051 respectively). MFR is significant in both the multivariable model (*p* < 0.0005) and stepwise backwards elimination model (*p* < 0.0005). Race is marginally insignificant in the multivariable model (*p* = 0.066) and significant in the backwards elimination model (*p* = 0.030). Repeating the partial multivariable model including these two predictors does not change the significance of any of the other predictors. In this case race remains insignificant (*p* = 0.188), but males per 100 females becomes significant (0.0106, 95% CI: 0.0060 to 0.0153, *p* < 0.0005). One additional male per 100 females leads to an increase in detected cases of 1.1% (95% CI: 0.6% to 1.6%, *p* < 0.0005) in the multivariable model, 1.1% (95% CI: 0.6% to 1.5%, *p* < 0.0005) in the adjusted partial multivariable model, and 1.2% (95% CI: 0.7% to 1.7%, *p* < 0.0005) in the stepwise backwards elimination model.

### Final model

A final model was built using males per 100 females, percentage of dependent population, and median household income as predictors. Table 3 shows a summary of the regression estimates from this model. Fig 7A shows the area specific relative risk *ζ*_*i*_ for this model, whilst Fig 7B shows the posterior probability that the relative risk is greater than 1, *p* (*ζ*_*i*_ *>* 1| **y**). In this model, a 1% increase in the dependent population leads to a 2.5% (95% CI: 1.6% to 3.4%, *p* < 0.0005) increase in detected COVID-19 cases. An increase of 1 male per 100 females leads to a 1.0% (95% CI: 0.6% to 1.5%, *p* < 0.0005) increase in detected cases. Finally, a $10,000 decrease in median household income leads to a 2.5% (95% CI: 1.0% to 4.1%, *p* = 0.002) increases in detected cases. Fig 8 shows the percentage of the population testing positive for COVID-19 against each of these three predictors, along with our regression estimates and CI.

**Table 3.** Regression estimates for final model of association of Zip Code Tabulation Area (ZCTA) level predictors with detected COVID-19 cases in New York City as at April 5, 2020.

| Predictors | Estimate | 95% CI | p-value |
| --- | --- | --- | --- |
| Dependents<sup>1</sup> | 0.0246 | 0.0157, 0.0334 | <0.0005* |
| MFR<sup>2</sup> | 0.0100 | 0.0055, 0.0144 | <0.0005* |
| Income<sup>3</sup> | -0.0025 | -0.0040, -0.0010 | 0.002* |

Notes:
<sup>1</sup>Percentage of population under 18 or 65+.
<sup>2</sup>Males per 100 females.
<sup>3</sup>Median household income in \$1,000s.

**Fig 7.**
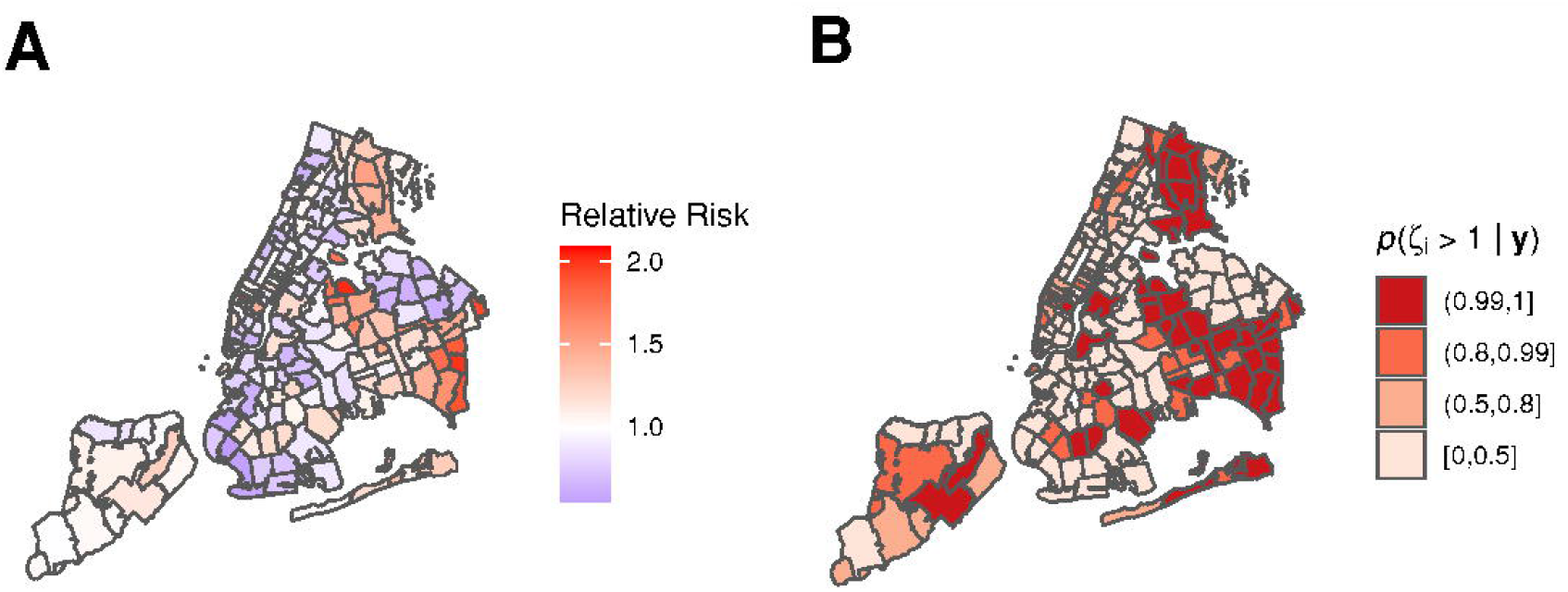
Ecological regression model for COVID-19 cases in New York City by Zip Code Tabulation Area (ZCTA). As at April 5, 2020, using final Poisson BYM2 model including percentage of dependent population, males per 100 females, and median household income as predictors. A: Area specific relative risk, *ζ*_*i*_. B: Posterior probability for relative risk, *p* (*ζ*_*i*_ *>* 1|**y**).

**Fig 8.**
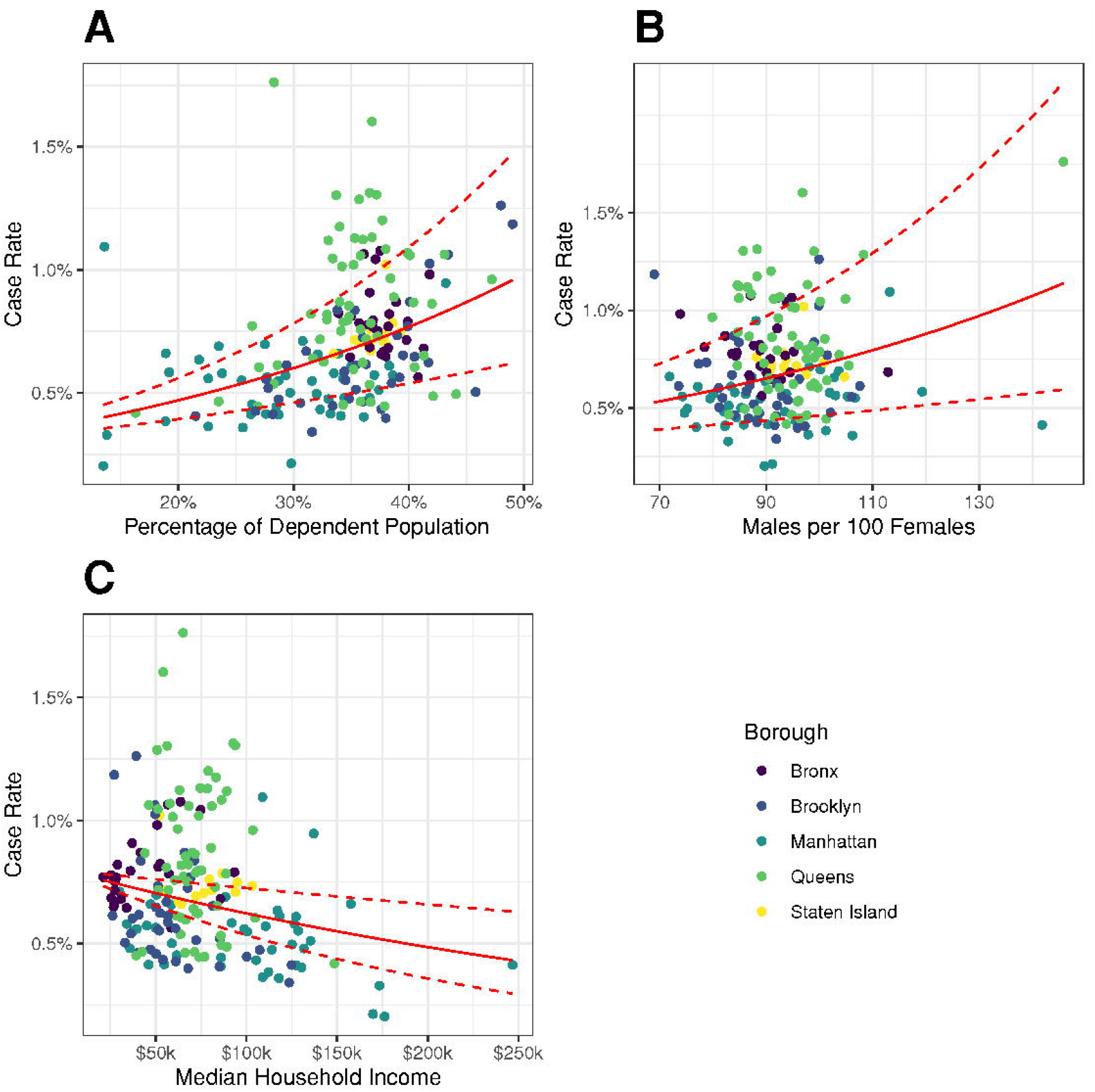
Percentage of population testing positive for COVID-19 in New York City by Zip Code Tabulation Area (ZCTA) against predictors used in final model. As at April 5, 2020, using final Poisson BYM2 model. Red regression lines show model estimates and 95% Confidence Interval (CI) with other predictors held at their mean values. A: Percentage of dependent population. B: Males per 100 females. C: Median Household Income.

## Discussion

During the opening stages of the COVID-19 pandemic in NYC, there has been considerable variation in detected cases between neighborhoods in the city. Disease mapping shown in Fig 5 displays a number of high risk areas, notably around Rikers Island, Southeast Queens, East Bronx, and the orthodox Jewish community around Borough Park, Brooklyn. The unprecedented national response has included large number of media stories touting various covariates as predictors of either COVID-19 cases or mortality. In this ecological study, we attempted to use spatial modeling techniques to assess the association between number of COVID-19 cases detected in different neighborhoods of NYC and neighborhood-level predictors. Our findings indicated a significant direct association between detected cases and both the proportion of dependents in the population and male to female ratio. We also see a significant inverse relationship between detected cases and median household income. We do not see a consistently significant relationship between detected cases and the other potential predictors; even those such as poverty, unemployment, and proximity to healthcare that were significant in a univariable model.

Our findings indicated statistically significant associations between two of the four demographic predictors included in the study. We find percentage of dependents in the population to be a statistically significant predictor in all of the models in which it appears as a factor. Conversely, we find that the aged percentage of the population (65+) is never a significant predictor of detected COVID-19 cases. We believe that taken together, this indicates that the youthful end of the dependent population is associated with higher case numbers. This is congruent with evidence from Chan et al. [14] and Bai et al. [15], both of whom suggest significant transmission by young asymptomatic carriers. We further hypothesize that attitudes and behavioral patterns could play a significant role in this effect. As an example, increasing mortality of COVID-19 with age has been well publicized, and we suggest this may incline older communities to adhere to preventative public-health measures more. Conversely the same information may be interpreted by younger populations that they are not at significant risk, potentially encouraging riskier behaviors.

The balance of males and females as a significant factor indicates that areas with more males are associated with higher detected COVID-19 cases. This factor remains significant even when the two outliers shown in Fig 8B, ZCTAs 10007 (downtown Manhattan) and 11370 (East Elmhurst including Rikers Island), are removed from the analysis. Wenham et al. [35] note the lack of sex analysis by global health institutions. Studies have posited sex differences in immunological function [36] or smoking prevalence/pattern [37] as potential causes of differing medical outcomes. We found no studies to date examining sex specific behavior trends in relation to COVID-19 transmission and incidence. Looking back further, we found conflicting evidence from studies on the 2009 H1N1 pandemic. Some studies suggested that females were more willing to engage in public health precautions [38], whilst others suggested no significant sex effects [39]. We suggest that further studies be undertaken to consider whether sex specific behavioral, employment, or other trends are mechanisms that could explain sex effects on detected cases. Race was marginally insignificant in our statistical analysis. Thus, we caution against any association beyond the confounding sociological relationship between race and economic affluence [40]. Further, our study used percentage of population identifying as white, splitting the demographic into two groups, white and non-white. A study exploring multiple racial categories at an aggregate and/or individual level, may draw additional insight [19].

Regarding the economic predictors, we note that our findings are in agreement with a previous, non-pandemic study [41], which found that affluence (in our case household income) was a significant predictor on self-rated health whilst poverty and income inequality (the Gini index) were not significant factors. Wen et al. suggest that the presence of affluence sustains neighborhood social organizations, which in turn positively affect health. If we extend this argument to the current pandemic, we could hypothesize that these social organizations further act to pass on information and promote community adoption of transmission-reduction policies such as social distancing [42].

Furthermore, we note that those in low affluence neighborhoods are more likely to live in higher density residence arrangements, for example community housing and shared family dwellings. The increased urban density likely contributes to transmission of the virus among the neighborhood [43]. Whilst previous studies [44] have found influence of unemployment on disease transmission, we note that the unprecedented shut-down of national infrastructure and the economy has meant that many previously employed people suddenly found themselves either unemployed, furloughed, or working from home. In a short period of time this drastic measure has completely altered the employment landscape of NYC such that it is unsurprising the predictor from 2018 is not significant.

We found that neither of our healthcare-related predictors was consistently significant. Lack of insurance has previously been a barrier to both diagnosis and treatment [45, 46]. However, in the COVID-19 pandemic, significant state resources were directed such that testing was freely available to all eligible New York residents.

We note four key limitations of the ecological study. First, our dependent variable is the number of detected COVID-19 cases, which may be significantly different from the number of true cases [47]. We believe, however that this does not detract from the validity of the study, since characterization of the detection and prevalance is important for pandemic management [48]. Studies on HIV rates amongst at risk populations suggest that the relationship between predictors and the number of detected cases is likely a complex interaction via at least three pathways: the true number of cases, access to testing (means) [49], and population attitudes to testing (motivation) [50, 51]. Thus, we can still develop valid inferences, even if we cannot elicit with certainty which one (or ones) of these pathways the significant predictors act through. Second, any associations made must be interpreted with caution since, as with any observational study, spurious correlations produced by unstudied confounding factors may be present. Caution is also advised due to the ecological fallacy of making individual inferences from aggregate data. Further verification is required to determine true causative links between predictors and detected cases even when associations are significant. Third, the significant predictors found are likely not the only explanations for different detection rates between different neighborhoods. However this study does provide useful insight into explaining between-neighborhood variation. Finally, since testing has been coordinated within the city limits at the borough level, there may be borough-level biases relating to COVID-19 testing. However, if these biases exist, they likely inhibit testing access in low-income neighborhoods [52, 53] such that the inverse association found between income and positive cases is more pronounced than what the model suggests.

## Conclusion

Within the constraints imposed by the limitations of an ecological analysis, we conclude that there exist consistent, significant associations between COVID-19 detected cases and the percentage of dependents in the population and male to female ratio. Further, there is also a significant association between detected COVID-19 cases and low income neighborhoods. The significance of dependents likely comes from the youth rather than the aged population. We suggest further studies be undertaken to determine any underlying causative mechanisms to these associations, paying particular attention to willingness to engage in public health behaviors and to asymptomatic carrier transmission. We finally highlight that whilst predictors may change with increased time and access to testing, this study provides important insights into public health behavior in the early stages of the current, and future, pandemic.

## Data Availability

The datasets analyzed for this study are publicly available, a repository can be found on GitHub.

https://github.com/rswhittle/NYC-COVID19-socioeconomic

## Acknowledgments

Figures were created using shapefiles publicly available from the NYC Geodatabase (NYC GDB) project [54].

## Author contributions

R.S.W. conceived and designed the work. R.S.W. collected data. R.S.W. designed the model and the computational framework and analysed the data. R.S.W. drafted the manuscript. R.S.W. and A.D.-A. revised the manuscript for critical intellectual content. R.S.W. and A.D.-A. approved the final version of the manuscript.

## Data availability statement

The datasets analyzed for this study are publicly available, a repository can be found on GitHub: https://github.com/rswhittle/NYC-COVID19-socioeconomic.

